# Validation of patient-reported outcome measures for dactylitis, psoriatic skin and nail disease, and uveitis in patients with psoriatic arthritis in routine care

**DOI:** 10.1101/2025.03.21.25324305

**Authors:** Louise Majormoen Nielung, Lene Wohlfahrt Dreyer, Lone Skov, Merete Lund Hetland, Bente Glintborg

## Abstract

**Background:** In routine care, Danish patients with psoriatic arthritis are monitored in the DANBIO registry. In March 2022, patient-reported outcome measures (PROMs) on selected non-musculoskeletal manifestations (NMM) were implemented.

**Aim:** To validate PROMs for current dactylitis, skin and nail psoriasis, and recent uveitis in patients with psoriatic arthritis.

**Methods:** Adaptive cross-sectional study. Patients in the rheumatologic clinic answered PROMs with “yes”/”no”/”do not know” and assessed extent of skin psoriasis and number of dactylitis-affected digits in DANBIO. PROM entries were compared to the physician’s assessments (physical examination, review of patient file), with physician being gold standard. With 134 patients included, 20% had incorrectly reported dactylitis; therefore, a dactylitis-photo was added to the PROM. Sensitivity, specificity, positive and negative predictive values, and accuracy were calculated. Level of agreement for dactylitis count was explored by Bland-Altman plot. From patient 200, the physician was blinded to PROs.

**Results:** We included 300 patients (51% female, median age=55 years), median disease duration: 8 years, 43% received biologic treatment. According to physician’s assessment, 41(14%) patients had current dactylitis, 164(55%) psoriasis, 163(54%) nail psoriasis, and 3(1%) recent uveitis. Agreement between patient and physician was high, with sensitivity/specificity for dactylitis 0.89/0.81, psoriasis 1.0/0.94, nail psoriasis 0.76/0.94, uveitis 1.00/0.99. Agreement on psoriasis-extent was 90%. Patient-reported dactylitis count was on average 1.0 unit higher than physician-reported but decreased to 0.7 after adding the dactylitis-photo.

Results were similar irrespective of blinding.

**Conclusion:** Patients reliably self-report dactylitis, psoriasis, and uveitis, and the PROMs are valuable for ruling out NMM in routine care.

## INTRODUCTION

Psoriatic arthritis is a chronic multifaceted inflammatory disease that involves the joints and the spine and is associated with non-musculoskeletal manifestations (NMMs), including psoriatic skin and nail disease, uveitis, and inflammatory bowel disease[1]. Furthermore, dactylitis and enthesitis are common manifestations[2]. If the treatment target is not reached with a conventional synthetic disease-modifying drug (csDMARD), the European Alliance of Associations for Rheumatology (EULAR) recommends that the choice of the biologic and targeted synthetic (b/ts-) DMARD should reflect whether NMMs related to psoriatic arthritis are present or not[3]. This highlights the importance of identifying and monitoring NMM in routine care to optimize treatment and improve patient outcomes[4].

Patient-reported outcomes (PROs) may be defined as “*any report coming directly from patients about a health condition and its treatment*”[5]. They are valuable for managing a range of chronic diseases[5–7], and in psoriatic arthritis, they improve patient-clinician communication and guide clinical decisions[8,9]. Currently, no validated and accepted PRO measures (PROMs) covering all aspects of NMM exist for patients with psoriatic arthritis, but a few studies have validated PROMs for selected NMM in psoriatic arthritis[10,11]. Thus, a PROM inspired by the Psoriatic Arthritis Disease Activity Score (PASDAS) has been developed[10]. It includes patient-reported psoriasis, dactylitis, enthesitis, and tender and swollen joint counts but has currently only been validated in patients with low disease activity[10].

Psoriasis Symptom Inventory (PSI), initially invented for patients with psoriasis, is a PROM for the severity of psoriasis symptoms, including itching, pain, redness, scaling, burning, cracking, and bleeding, which was found to correlate well with physician-reported body surface area (BSA) in patients with Psoriatic arthritis and mild psoriasis[11,12]. In patients with psoriasis, various PROMs regarding dactylitis, joint pain, skin and nail psoriasis are used in dermatology as part of screening tools for psoriatic arthritis[13–17]. Focus has been on validating their ability to detect psoriatic arthritis rather than on the validity of detecting the NMM themselves[13–17].

In the Danish nationwide rheumatology registry, DANBIO, >15,000 patients with psoriatic arthritis have been monitored prospectively in routine care[18]. Until recently, focus has been on joint manifestations, whereas information on NMM has not routinely been recorded. To address the multifaceted nature of psoriatic arthritis, new PROMs were introduced in March 2022, covering current dactylitis, skin and nail psoriasis, and recent uveitis. We hypothesized that patients with psoriatic arthritis can accurately self-report these manifestations. This study, using an adaptive study design, aimed to improve and validate these PROMs in patients with psoriatic arthritis.

## METHODS

This adaptive cross-sectional monocentric study was conducted between December 2023 and May 2024 at the outpatient clinic at the Department of Rheumatology, Copenhagen University Hospital, Rigshospitalet Glostrup, where more than 1,000 patients with psoriatic arthritis are followed. All patients with a rheumatologist-confirmed diagnosis of M07.3A (psoriatic arthritis), M07.3B (psoriatic arthropathy), or M46.8+M07.3 (inflammatory spondylopathies with psoriatic arthritis) were consecutively invited to participate when they attended the outpatient clinic for a scheduled consultation. Patients were included if they spoke Danish, could comply with the examination program, and had completed the NNM PROMs no more than seven days before the consultation. Newly diagnosed patients (< 3 months) were excluded.

**Table 1** shows the PROMs for dactylitis, psoriatic skin and nail disease, and uveitis, here termed NMM PROMs. The PROMs addressed whether the patients had current dactylitis, current psoriatic skin and nail disease and whether they had had uveitis since the last consultation. The patients filled out the NMM PROMs in DANBIO before the consultation (at home, via www.danbio.dk, or by touch screens in the clinic[19]). The PROMs were considered self-explanatory, and no patient education was provided. In addition, the patients filled out routine PROMs for psoriatic arthritis in DANBIO (patient pain, fatigue, and global assessment on Visual Analogue Scales (VAS), the Multi-Dimensional Health Assessment Questionnaire (MDHAQ), and Patient Acceptable Symptom State (PASS))[18].

**Table 1.**
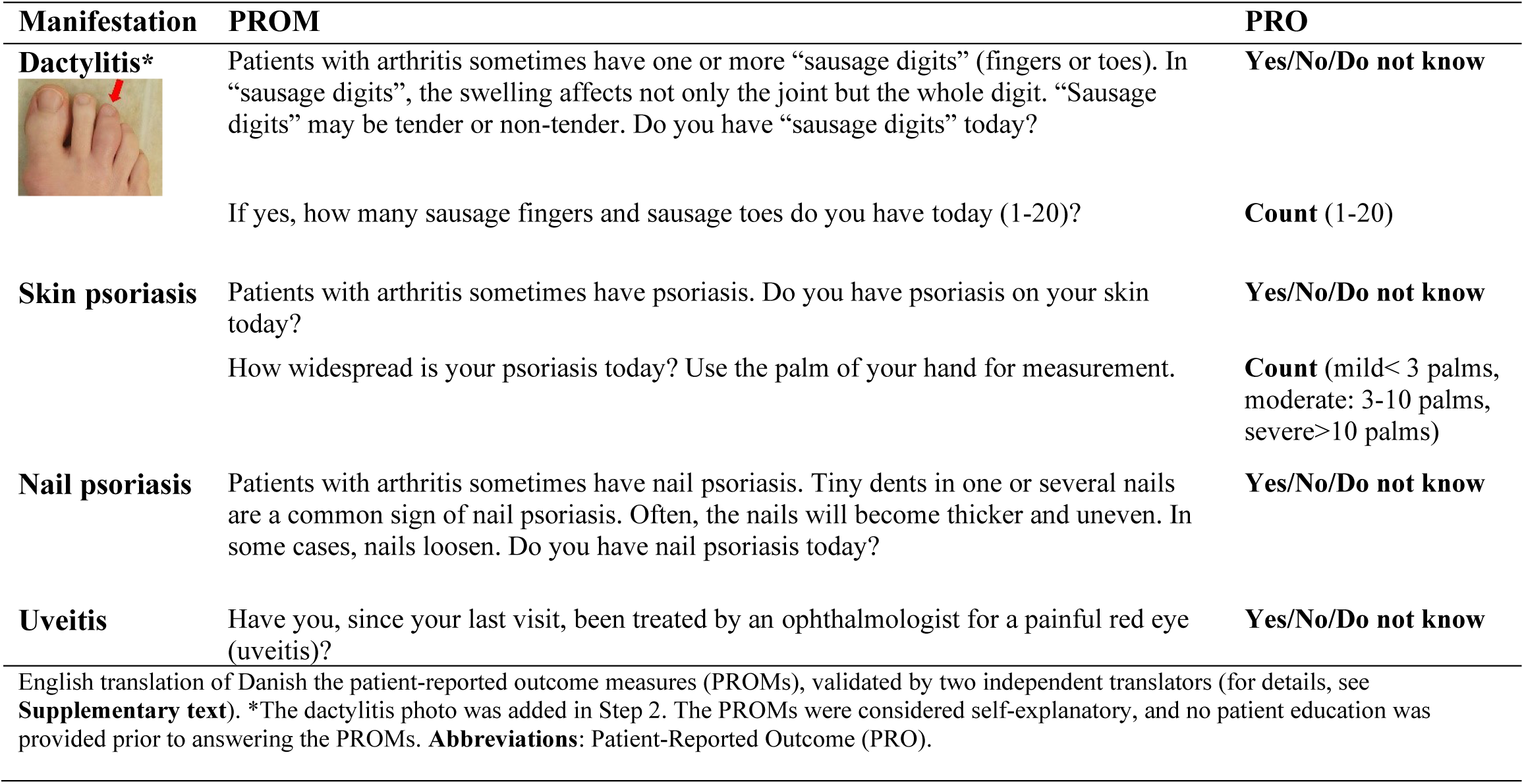
Patient-Reported Outcome Measures for non-musculoskeletal manifestations in DANBIO.

| Manifestation | PROM | PRO |
| --- | --- | --- |
| **Dactylitis\*** | Patients with arthritis sometimes have one or more “sausage digits” (fingers or toes). In “sausage digits”, the swelling affects not only the joint but the whole digit. “Sausage digits” may be tender or non-tender. Do you have “sausage digits” today?<br>If yes, how many sausage fingers and sausage toes do you have today (1-20)? | **Yes/No/Do not know**<br>**Count** (1-20) |
| **Skin psoriasis** | Patients with arthritis sometimes have psoriasis. Do you have psoriasis on your skin today?<br>How widespread is your psoriasis today? Use the palm of your hand for measurement. | **Yes/No/Do not know**<br>**Count** (mild< 3 palms, moderate: 3-10 palms, severe>10 palms) |
| **Nail psoriasis** | Patients with arthritis sometimes have nail psoriasis. Tiny dents in one or several nails are a common sign of nail psoriasis. Often, the nails will become thicker and uneven. In some cases, nails loosen. Do you have nail psoriasis today? | **Yes/No/Do not know** |
| **Uveitis** | Have you, since your last visit, been treated by an ophthalmologist for a painful red eye (uveitis)? | **Yes/No/Do not know** |

English translation of Danish the patient-reported outcome measures (PROMs), validated by two independent translators (for details, see **Supplementary text**). \*The dactylitis photo was added in Step 2. The PROMs were considered self-explanatory, and no patient education was provided prior to answering the PROMs. **Abbreviations:** Patient-Reported Outcome (PRO).

At the routine consultation, the rheumatologist in the outpatient clinic entered tender and swollen joint counts (28 joints) and current treatment into DANBIO. C-reactive protein was automatically imported from the laboratory database to calculate Disease Activity index in PSoriatic Arthritis based on 28 joints (DAPSA28)[20]. After the consultation, the patients were examined by a study physician (LN) who scored dactylitis, psoriatic skin, and nail disease, and uveitis (for details, see **Supplementary text**). The PROs and the study physician’s assessments were compared for validation, the latter being the gold standard. Both the patients and the study physician had the option to answer “yes”, “no”, or “do not know” as to whether the individual NNM was present.

The study design was iterative (**Figure 1**)[21]. Challenges that evolved during the study were discussed in the study group, leading to adaptations of the PROMs or the validation process, here termed adaptive steps. In all patients, fingers, toes, and nails were inspected for dactylitis and psoriatic nail disease. Dactylitis was assessed and scored using the “Leeds dactylitis index basic,” where a digit with dactylitis should have a digital circumference ≥ 10% compared to the contralateral (and nonaffected) digit (see **Supplementary text**)[22]. The assessment of uveitis (i.e., patient interviews validated by medical records and medication lists) and psoriatic nail disease remained unchanged during the study, whereas the assessment of dactylitis and psoriatic skin disease had the following three adaptive steps **(Figure 1)**:

**Step 1** (patient number 1 to 134): The study physician was not blinded to the PROs in order to simulate a clinical setting. The skin assessment for psoriatic lesions was guided by PROs and patient interviews. If the patient answered “no” or “do not know,” the study physician asked whether psoriasis was present in the different body parts to make sure no areas were overlooked. A single skin lesion smaller than 2 cm was not considered to be psoriasis.

**Step 2** (patient number 135 to 200): The study physician was not blinded to the PROs. A dactylitis photo was added to the dactylitis PROM following the observation that, among the first 134 patients, 20% had inaccurately reported the presence of dactylitis. The skin psoriasis evaluation was as described in Step 1.

**Step 3** (patient number 201 to 300): The study physician was blinded to the PROs. The skin psoriasis examination was done after removing all the patient’s clothing to examine the entire body. The dactylitis evaluation was as described in Step 2.

**Figure 1.**
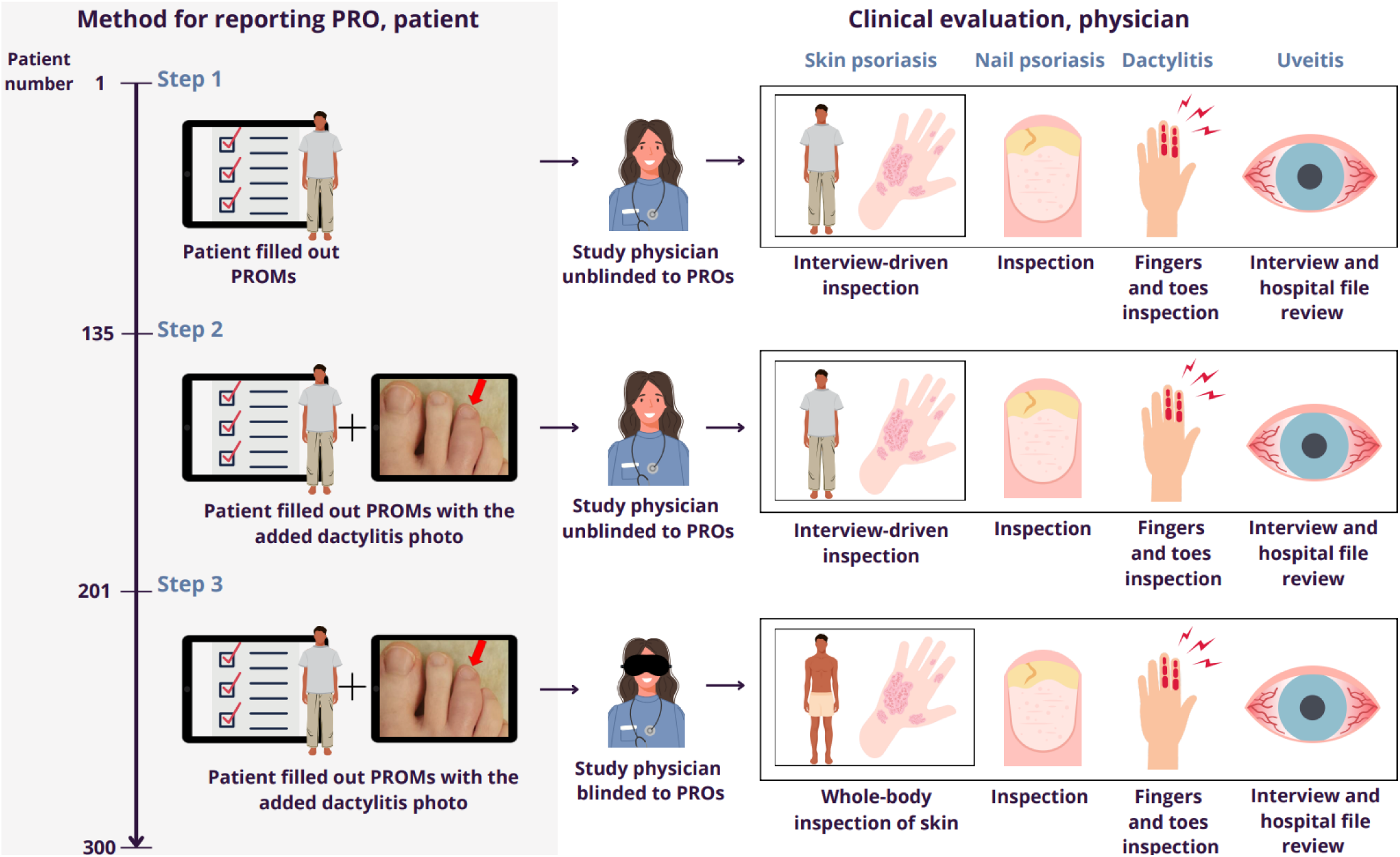
Study design. The inclusion and validation process in three adaptive steps. <u>Step 1 (patient number 1 to 134):</u> The patients filled out the NMM PROMs in DANBIO online (on the patient’s smartphone, tablet, or PC) or on a screen in the waiting area before consultations in the outpatient clinic[19]. The physician was not blinded to the PROs to simulate a clinical setting. For individual patients, skin inspection for psoriatic lesions was guided by PRO response and patient interview. In all patients, fingers, toes, and nails were inspected for psoriatic nail disease and dactylitis[22]. Uveitis was validated by interviews and medical records including medication lists. The evaluation for uveitis and psoriatic nail disease remained unchanged during the study. <u>Step 2 (patient number 135 to 200)</u>: A dactylitis photo was added to the dactylitis PROM to reduce false positive responses. The physician was not blinded to the PROs. The clinical evaluation was as described in Step 1. <u>Step 3 (patient number 201 to 300):</u> The physician was *blinded* to the PROs to avoid confirmation bias. Psoriatic skin disease was evaluated after removing all the patients’ clothing to examine the entire body, irrespective of the patient’s self-report. The clinical evaluation for psoriatic nail disease, dactylitis, and uveitis was as described in Step 1. **Abbreviations:** Patient-reported outcome measure (PROM). Patient-reported outcome (PRO). Non-musculoskeletal manifestations (NMM).

### Sample size estimation

The statistical power was calculated based on literature estimates of the prevalence of current NMM: 40% for dactylitis, 60% for psoriasis, and 60% for psoriatic nail disease[23]. With a sample size of 300 patients, the estimated precision was ±5.0%, with a confidence level of 0.9 for dactylitis, psoriatic skin disease, and nail disease[24,25]. The expected prevalence of uveitis was 3%[23], resulting in too few expected cases to fully validate this manifestation when including 300 patients[25,26].

### Statistical methods

Sensitivity, specificity, positive predictive value, negative predictive value, and accuracy were calculated for all categorical PROMs. The level of agreement between the patient’s and study physician’s dactylitis count was visualized with Bland-Altman plots. The extent of psoriasis was measured as body surface area (BSA) in three categories: mild: <3 palms, moderate: 3-10 palms, and severe: >10 palms[27]. The level of agreement between the patient and study physician was calculated as the number of times the study physician and patient gave the same assessment, expressed as a percentage of the total number of assessments.

The data used for validation were complete with no missingness. The disease characteristics obtained from DANBIO, presented in Table 2, were not imputed if missing.

**Table 2.**
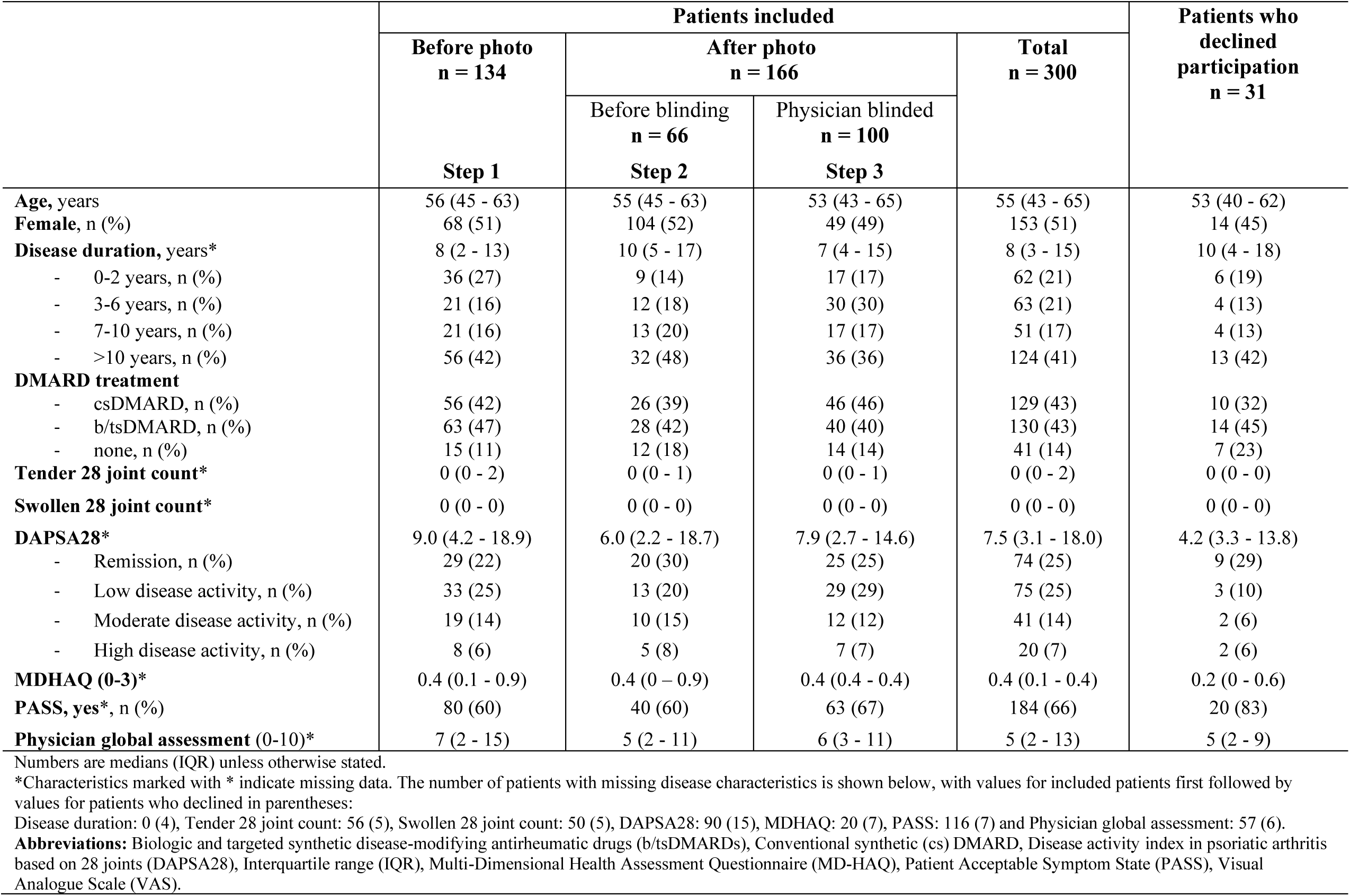
Demographics and disease characteristics of the included patients (stratified by the three adaptive steps), and of the patients who declined participation.

### Ethics

Patients were included after written informed consent. The Ethics Committee renounced study approval (F-23061081) because of the non-interventional study design. Data collection was handled according to the General Data Protection Regulation (EU) 2016/679 and approved by the Capital Region of Denmark (p-2023-14943). Results are reported according to the STARD guidelines [28].

## RESULTS

### Study population

We enrolled 300 patients with psoriatic arthritis with a median age of 55 years (IQR 43-65 years), of whom 51% were female (**Table 2**). Most, but not all, patients had longstanding disease, and the median disease duration was 8 years (IQR 3-15 years). In 43% of the patients, the current treatment was conventional synthetic disease modifying drug (csDMARDs), 43% received biologic and targeted synthetic (b/ts)DMARD (as monotherapy or in combination with csDMARD), and 13% received no DMARD. The median 28 tender and swollen joint counts were 0, and the median disease activity (DAPSA28) was 7.5 (25% were in remission, and 25%, 14%, and 7% had low, moderate, and high disease activity, respectively)[29]. A total of 31 patients declined participation, primarily due to lack of time (**Supplementary Figure S1**). Patients who declined were overall younger, more often male, had longer disease duration, were less likely to receive DMARD therapy, had less disability, and more often scored PASS=yes compared to included patients (**Table 2**).

At the physical examination, the study physician found that 41 patients (14%) had current dactylitis, 164 (55%) psoriasis (91% mild, 8% moderate, 1% severe), 163 (54%) psoriatic nail disease, and 3 (1%) had had uveitis since the last visit. The median dactylitis count was 0 (0-0). Among the patients with dactylitis, the median dactylitis count was 2 (1-3).

### Validation results

#### Dactylitis

The sensitivity for the dactylitis PROM was 0.75 before we added the dactylitis photo (Step 1) and increased to 0.88 after the photo had been added (Steps 2+3). Corresponding specificities were 0.79 before (Step 1) and 0.83 after (Steps 2+3). The specificity was independent of whether the study physician was unblinded or blinded to the PROs (0.88 and 0.81, respectively). The corresponding sensitivity was 0.88 and 0.89, respectively (**Table 3**). The patients often confused dactylitis with other conditions, including synovitis, osteoarthritis, and edema.

**Table 3.** Sensitivity, specificity, positive and negative predictive value, and accuracy of the non-musculoskeletal manifestation PROMs in Step 1, 2, and 3.

| Current manifestation | Step | N | PRO<br>yes/no/do<br>not know* | Study<br>physician<br>n<br>yes/no/do<br>not know* | Sensitivity<br>(95% CI) | Specificity<br>(95% CI) | PPV<br>(95% CI) | NPV<br>(95% CI) | Accuracy<br>(95% CI) |
| --- | --- | --- | --- | --- | --- | --- | --- | --- | --- |
| **Dactylitis** |  |  |  |  |  |  |  |  |  |
| -without photo | 1 | 134 | 35/91/8 | 13/120/1 | 0.75<br>(0.43, 0.95) | 0.79<br>(0.70, 0.86) | 0.26<br>(0.13, 0.44) | 0.97<br>(0.91, 0.99) | 0.78<br>(0.69, 0.85) |
| -after photo | 2+3 | 166 | 43/104/19 | 28/138/0 | 0.88<br>(0.70, 0.98) | 0.83<br>(0.76, 0.90) | 0.53<br>(0.38, 0.69) | 0.97<br>(0.92, 0.99) | 0.84<br>(0.77, 0.90) |
| -before blinding of study physician | 2 | 66 | 14/46/6 | 9/57/0 | 0.88<br>(0.47, 1.00) | 0.88<br>(0.77, 0.96) | 0.54<br>(0.25, 0.81) | 0.98<br>(0.89, 1.00) | 0.87<br>(0.75, 0.94) |
| -after blinding of study physician | 3 | 100 | 29/58/13 | 19/81/0 | 0.89<br>(0.65, 0.99) | 0.81<br>(0.70, 0.90) | 0.55<br>(0.36, 0.74) | 0.97<br>(0.88, 1.00) | 0.83<br>(0.73, 0.90) |
| **Skin psoriasis** |  |  |  |  |  |  |  |  |  |
| -before blinding | 1+2 | 200 | 115/81/4 | 110/88/2 | 0.99<br>(0.95, 1.00) | 0.95<br>(0.88, 0.99) | 0.96<br>(0.91, 0.99) | 0.99<br>(0.93, 1.00) | 0.97<br>(0.94, 0.99) |
| -after blinding of study physician | 3 | 100 | 59/32/9 | 54/40/6 | 1.00<br>(0.93, 1.00) | 0.94<br>(0.80, 0.99) | 0.96<br>(0.88, 1.00) | 1.00<br>(0.89, 1.00) | 0.98<br>(0.92, 1.00) |
| **Psoriatic nail disease** |  |  |  |  |  |  |  |  |  |
| -before blinding | 1+2 | 200 | 82/101/17 | 101/98/1 | 0.82<br>(0.73, 0.89) | 0.95<br>(0.89, 0.99) | 0.95<br>(0.88, 0.99) | 0.83<br>(0.74, 0.90) | 0.88<br>(0.82, 0.92) |
| -after blinding of study physician | 3 | 100 | 47/49/4 | 62/36/2 | 0.76<br>(0.63, 0.86) | 0.94<br>(0.81, 0.99) | 0.96<br>(0.85, 0.99) | 0.71<br>(0.56, 0.83) | 0.83<br>(0.74, 0.90) |
| **Uveitis** | 1-3 | 300 | 7/292/1 | 3/297/0 | 1.00<br>(0.29, 1.00) | 0.99<br>(0.97, 1.00) | 0.43<br>(0.10, 0.82) | 1.00<br>(0.99, 1.00) | 0.99<br>(0.97, 1.00) |

Sensitivity: Proportion of patients answering “yes” to manifestation and having it according to study physician evaluation. Specificity: Proportion of patients answering “no” to manifestation and not having it. Positive predictive value (PPV): Probability for a yes-answer to be true. Negative predictive value (NPV): Probability for a no-answer to be true. \*The ‘do not know’ answers from the study physician and patients were excluded from sensitivity, specificity, PPV, NPV, and accuracy analyses. **Abbreviations:** Number of patients (N), patient-reported outcome measure (PROM), 95 % confidence interval (95% CI).

The differences between the patient-reported and physician-reported dactylitis count after we added a dactylitis photo (Step 2+3) are shown as Bland-Altman plot (**Figure 2**). The patient-reported dactylitis count was on average 1.0 (95% CI: -5.4 to 7.4) units higher than the study physician’s but decreased to 0.7 (95% CI -3.7 to 5.2) after adding the dactylitis photo. In patients with dactylitis (N=41), the patients and study physician agreed on the dactylitis count in 14 of 41 (34 %), whereas 17 (41%) patients overrated, 7 (17%) underrated the dactylitis count, and 3 (7%) patients answered, “do not know”. In three cases, patients reported having a dactylitis count of 20 while the study physician reported 0 (one case was after the dactylitis photo was added).

**Figure 2.**
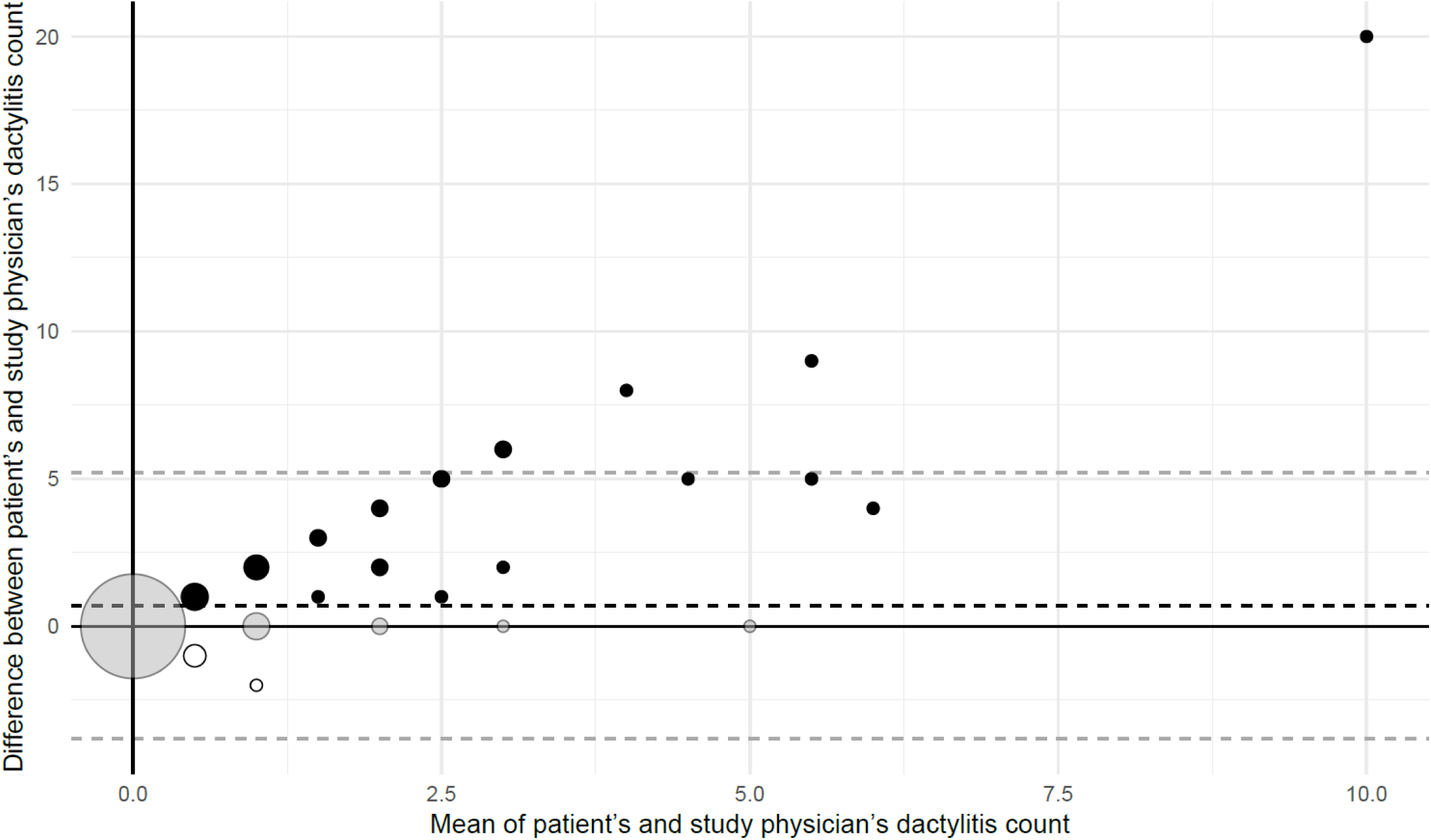
Bland-Altman Plot of dactylitis count. Bland-Altman plot of the difference (patient’s count minus study physician’s count) *after* adding a dactylitis photo (Step 2+3) to the PROM. The bold dotted black line is the average difference between the patient’s and the study physician’s dactylitis count (0.7). The dotted lower (- 3.7) and upper (5.2) grey lines are the lower and upper 95% confidence interval lines, respectively. Grey dots: The patient and study physician agreed on the dactylitis count. Black dots: The patient scored higher than the study physician. White dots with black border: The patient scored higher than the study physician. The dot size reflects the number of patients.

The results from Table 3 presented as crosstabulation according to STARD guidelines can be found in **Supplementary Table S1**[28]. Positive predictive value (PPV), negative predictive value (NPV), and accuracy for each NMM are shown in Table 3.

#### Skin psoriasis

The sensitivity of the skin psoriasis PROM did not change when the study physician in Step 3 was blinded to the PROs and did a whole-body examination for skin psoriasis (0.99 and 1.0, respectively). The corresponding specificities were also similar (0.95 and 0.94) (**Table 3**). The level of agreement on the presence and extent of psoriasis was 90% (**Table 4**). Few patients answered, “do not know,” and those who did, in general, did not have psoriasis. In seven cases, the study physician could not determine if the patient had skin psoriasis – in three of them, the patient answered, “do not know”.

**Table 4.** The level of agreement on the presence and extent of psoriasis (Step 1-3) between patient and study physician.

|  |  | Study physician assessment |  |  |  |  |
| --- | --- | --- | --- | --- | --- | --- |
| PRO |  | Not present | Mild | Moderate | Severe | Do not know |
|  | Not present | 113 | 1 | 0 | 0 | 0 |
|  | Mild | 5 | 132 | 0 | 0 | 4 |
|  | Moderate | 1 | 17 | 12 | 1 | 0 |
|  | Severe | 0 | 0 | 1 | 0 | 0 |
|  | Do not know | 10 | 0 | 0 | 0 | 3 |

The agreement of the presence and extent of psoriasis was measured as body surface area (BSA) in three categories: mild<3 palms, moderate: 3-10 palms, and severe>10 palms. The grey-scaled areas are the cases where the patient and the study physician agree on the presence and extent of psoriasis.
**Abbreviations:** Patient-reported outcome (PRO).

#### Psoriatic nail disease

The sensitivity for nail psoriasis PROM was 0.82 and 0.76, respectively, before and after the study physician was blinded to the PROs, and the specificity was unchanged (0.95 and 0.94, respectively) (**Table 3**).

#### Uveitis

The sensitivity for the uveitis PROM was estimated to be 1.0, and the specificity to be 0.99 (**Table 3**).

## DISCUSSION

In this study, we investigated the validity of PROMs for dactylitis, psoriatic skin and -nail disease, and uveitis in 300 patients with psoriatic arthritis and demonstrated good agreement between the patient’s and study physician’s evaluation.

PROs improve communication between patients and healthcare providers and allow patients to monitor their disease and engage in self-management[6]. Patients’ self-report enables systematic collection of information independently of physicians, and it fosters a patient-centered focus during consultations[30].

To our knowledge, only a few clinical rheumatologic registries collect PROs on dactylitis, skin and nail psoriasis, and uveitis in routine care[31]. A previous study describing the set-up of 15 European registries within spondyloarthritis reported the collection of dactylitis, skin and nail psoriasis, and uveitis[32] However, the mode of assessment (physician or patient-reported) was not described. A systematic review assessed the outcome measures collected in 27 ongoing Psoriatic arthritis registries or longitudinal cohorts[33]. All items were physician-reported and included skin psoriasis in 93% of the registries (PASI 4 in 66%, BSA in 44%) and dactylitis in 74% of the registries[33]. Outcome measures assessing nail psoriasis and uveitis were not collected since they are not part of the OMERACT/GRAPPA psoriatic arthritis core domains[34]

In some patients with psoriatic arthritis, physicians may find dactylitis challenging to assess, and multiple approaches can be used[2,35]. In the current study, dactylitis was the most challenging manifestation for patients to report accurately, with a tendency to overreport. Our patients often confused dactylitis with other conditions, including synovitis, osteoarthritis, and edema, but sensitivity and specificity improved after we added the dactylitis photo to the PROM. A similar tendency for patients with psoriatic arthritis to overreport dactylitis count has been observed in other studies, particularly in patients with low disease activity, with reported differences in dactylitis score ranging from 0 to 11[10]. The dactylitis prevalence in our study (14%) was lower than what has been reported in previous studies (15-50%), reflecting that this was a well-treated patient group, of whom most, but not all, had established disease[36–38].

While rheumatologists agree on the importance of psoriatic skin assessment in psoriatic arthritis, patient self-reporting has not yet been standardized[11]. The Psoriasis Symptom Inventory (PSI), a PROM, measures how the psoriasis symptoms affect the patient, but does not include a direct assessment of the extent of psoriasis[12]. A study showed that PSI correlates well with physician-reported BSA in patients with psoriatic arthritis[11]. A Self-Administered version of Psoriasis Area Severity Index (SA-PASI) has shown a significant correlation (Spearman’s correlation coefficient = 0.70) with PASI, though patients consistently report higher scores than physicians[39,40]. In the current study, the study physician was blinded to the PROs to investigate potential confirmation bias. It did, reassuringly, not markedly change the results, and the added whole-body examination for psoriasis did not reveal more psoriasis. These results support that BSA may be a suitable measure for self-reporting skin psoriasis in patients with psoriatic arthritis[10].

We found slightly more than half of the patients to have psoriatic nail disease (54%), which is in agreement with what has been reported by others[41,42]. As with dactylitis, the patients tended to overreport psoriatic nail disease, typically when they had longitudinal ridges in the nail. We observed that the study physician tended somewhat to align with the patient’s evaluation of nail psoriasis (confirmation bias), i.e., the sensitivity tended to be higher when the study physician was unblinded (0.82 (0.73-0.89)) than when blinded (0.76 (0.63-0.86)). That is an interesting finding, which, to our knowledge, has not yet been investigated by others.

As expected, uveitis was a rare manifestation. We found high sensitivity and specificity, but based on few events (n=3, 1%). Others have reported similar prevalence[43]. In a previous cross-sectional study in patients with spondyloarthropathies, 82% of the patients self-reported recent uveitis correctly[44]. In that study, the diagnosis was confirmed by a uveitis specialist reviewing ophthalmology records. In our study, we did not have access to records from private ophthalmologists. However, we expect this to be of minimal significance as cases of uveitis would be captured during the review of medication lists.

The PPV and NPV are affected by the prevalence of the disease and can differ from one setting to another. For skin and nail psoriasis, PPVs were high (close to 1), supporting that in our population these PROMs were performing very well compared to the gold standard.

The NPV was very high for dactylitis, skin and uveitis. The NPV predicts how likely it is for the patient to truly not have the NMM in case of a negative test result.

We estimated the accuracy of the PROMS by calculating the proportion of true positives and true negatives in all evaluated cases. Accuracy was high overall, highest for skin psoriasis and uveitis, lowest for dactylitis.

Our study was designed to reflect and accommodate the clinical situation in routine care. The study has several strengths. The patients were included during all clinic hours throughout the week resulting in practically consecutive recruitment, and few patients declined to participate. This limits the risk of selection bias. One study physician was responsible for recruitment and inclusion and conducted all examinations. The participating patients covered different disease durations and disease activity states. It increases the clinical feasibility of the PROMs that they were self-explanatory, and that no specific patient education was needed. The adaptive design allowed us to add a dactylitis photo, which reduced the patients’ tendency to overreport dactylitis, i.e., it led to increased sensitivity and, to a lesser extent, increased specificity[21].

Our study also has limitations. It was conducted at a university hospital which may result in selection bias due to lower representation of patients with milder disease who are typically treated in primary care. Furthermore, the number of patients with severe skin psoriasis was limited, so the PROMs’ performance in this subgroup remains less clear. We assessed dactylitis based on the clinical presentation. If ultrasound had been applied as the gold standard, this might have impacted the sensitivity and specificity of the dactylitis PROM. However, in many settings, ultrasound is not an option in routine care. Therefore, we deliberately excluded ultrasound from this study and chose the Leeds Dactylitis Index (LDI) basic, which is a feasible and validated alternative[22]. We were aware that our sample size of 300 patients was not sufficient to do a proper validation of the uveitis PROM because of its low prevalence. Therefore, the results for uveitis should be interpreted with caution. It may be seen as a weakness that we did not cover all extra-articular manifestations and NMM. However, this was a deliberate choice, as we considered enthesitis, axial involvement, and inflammatory bowel disease to be too complex for patients to self-report, consistent with the approach taken by most other registries[31].

Our findings may have implications for clinical care. Systematic monitoring of NMMs in psoriatic arthritis has the potential to provide important information on domains that often go unnoticed in a busy clinical setting[6]. By integrating NMM PROMs into routine care, a more comprehensive understanding of the patient’s condition can be achieved. This can not only enhance the patient’s self-management of the disease but also provide healthcare providers with valuable insights that can inform and optimize treatment strategies[6]. Our results showed that a negative response to the PROM generally ruled out the manifestation (high sensitivities, i.e. dactylitis, skin psoriasis, and uveitis, but not nail psoriasis).

With high specificities (skin and nail psoriasis, uveitis, but not dactylitis), a positive reply often confirmed the manifestation, and the physician should take the finding into account regarding, e.g., choice of b/ts DMARD treatment[3].

In conclusion, this adaptive cross-sectional validation study in 300 patients with psoriatic arthritis demonstrated that patients reliably self-report dactylitis, skin- and nail psoriasis, and uveitis. We found very high sensitivity and specificity for the skin psoriasis and uveitis PROMs, while we observed lower sensitivity for the dactylitis PROM and lower specificity for the nail psoriasis PROM. The PROMS are especially valuable for ruling out disease manifestations and for monitoring non-musculoskeletal manifestations in routine care.

## Data Availability

All data produced in the present study are available upon reasonable request to the authors

## ACKNOWLEDGMENTS

To patient representative Hans Albert Frandsen.

Dactylitis image courtesy of Dr. Ana-Maria Orbai, Director Johns Hopkins Psoriatic Arthritis Program, Baltimore, Maryland, USA

## COMPETING INTERESTS

LMN:

Grants or contracts from any entity: The Danish Rheumatism Association (paid to institution)

LD:

Grants or contracts from any entity: Contract with BMS and AbbVie outside the present work (paid to institution)

Support for attending meetings and/or travel: Janssen, UCB, Boehringer Ingelheim

Leadership or fiduciary role in other board, society, committee or advocacy group, paid or unpaid: Chair of the scientific committeé of the Danish Rheumatism Association (unpaid) and member of DANBIOS steering committee (unpaid)

LS:

Grants or contracts from any entity: Almirall, Janssen, Bristol-Myers Squibb, UCB, and Sanofi (paid to institution)

Payment or honoraria for lectures, presentations, speakers bureaus, manuscript writing or educational events: BMS, LEO Pharma, Pfizer, Janssen, AbbVie, Takeda and Sanofi (payment to LS)

Participation on a Data Safety Monitoring Board or Advisory Board: AbbVie, Novartis, Almirall, LEO

Pharma, Pfizer, Bristol-Myers Squibb, Boehringer Ingelheim, UCB, Eli Lilly, Stada, Takeda and Sanofi (payment to LS)

Leadership or fiduciary role in other board, society, committee or advocacy group, paid or unpaid: IPC, board member (unpaid)

MLH:

Grants or contracts from any entity: AbbVie, AlfaSigma, BMS, Eli Lilly, MSD, Pfizer, Sandoz, Novartis, Nordforsk, UCB (paid to my institution)

Payment or honoraria for lectures, presentations, speakers bureaus, manuscript writing or educational events: Pfizer, Medac, Sandoz, UCB (paid to institution) Novartis (paid to institution and to MH)

Participation on a Data Safety Monitoring Board or Advisory Board: AbbVie (paid to institution)

Leadership or fiduciary role in other board, society, committee or advocacy group, paid or unpaid: MLH has chaired the steering committee of the Danish Rheumatology Quality Registry (DANBIO, DRQ), which receives public funding from the hospital owners and funding from pharmaceutical companies. MLH co-chairs EuroSpA, which generates real-world evidence of treatment of psoriatic arthritis and axial spondylarthritis based on secondary data and is partly funded by Novartis and UCB.

BG:

Grants or contracts from any entity: Pfizer, AbbVie, Sandoz, Eli Lilly, AlfaSigma (paid to institution)

Leadership or fiduciary role in other board, society, committee or advocacy group, paid or unpaid: Chair of the DANBIO steering Commitee

