## Supplementary text, Supplementary Table S1, Supplementary Figure S1 for "Validation of patient-reported outcome measures for dactylitis, psoriatic skin and nail disease, and uveitis in patients with psoriatic arthritis in routine care"

METHODS. CLINICAL EVALUATION, DETAILED DESCRIPTION

All evaluations were conducted by the study physician, who assessed each manifestation as present (yes), not present (no), or unknown (do not know).

**Dactylitis:**

The clinical examination of dactylitis assessed the presence and graduation of dactylitis in fingers and toes (0-20). Using the “Leeds Dactylitis Index Basic”, a finger with dactylitis was defined by having a digital circumference ≥ 10% compared to the contralateral (and nonaffected) finger[1]. With a measuring tape, the circumferences of the the proximal phalanx of the affected fingers suspicious of dactylitis were measured, as close as possible to the interdigital fold. Circumferences of the contralateral fingers were measured at the same level. If both sides were affected, the circumference of the affected finger was compared to normative data[1]. The clinician squeezed the affected fingers with moderate pressure (enough to blanch the examiner’s nailbed)[1].

**Uveitis:**

Uveitis is rarely expected to be present at the visit; thus, questioning for uveitis symptoms and assessment of medical records was performed. The patients were interviewed regarding symptoms (pain, sensitivity to light, red eye, reduced vision) and treatment, if any. Medical records were searched for “uveitis,” and records from hospital ophthalmologists were screened, looking for ICD-10 codes (H20.011, H20.012, H20.013, H20.021, H20.22, H20.023, H20.11, H20.12, H20.13). Finally, medical history was collected in medication lists regarding uveitis treatment (Maxidex, Ultracortenol, Monopex).

**Skin psoriasis:**

At the clinical examination, the study physician asked whether the patient had current and/or previous skin and nail psoriasis and whether a dermatologist had verified the diagnosis. Additionally, the patients were asked about genetic predisposition for psoriasis. In case of doubt about the presence or extent of psoriasis, we had the opportunity to send photos to a consultant in dermatology for consultation. If the patient had never been to a dermatologist and/or the affected area was less than 2 cm, the study physician could not determine if the patient had psoriasis.

Step 1-2: A patient's skin was only examined and assessed if the patient answered "yes" to having current psoriasis in the skin, and the examination was guided by patient-interview.

If the patient answered "no" or "do not know," the physician asked whether psoriasis was present in the different body parts (arms, legs, torso, genitally, in and behind ears and at the scalp) and inspected the areas to which the patient answered "yes". Step 3: The skin psoriasis examination was done by removing all the patients' clothing to examine the entire body irrespective of the patient’s self-report.

The body surface area (BSA) affected by skin psoriasis was assessed to estimate the extent of psoriasis. The hand surface area is commonly used to assess BSA with an assumption that a hand surface represents 1% of the BSA[2]. Mild psoriasis affects less than 3% of the BSA, moderate psoriasis affects 3%-10%, and severe psoriasis affects more than 10% of the BSA[3].

**Nail psoriasis:**

The nails of hands and feet were evaluated for psoriatic changes, including onycholysis, pitting, and hyperkeratosis[4,5]. The nails of the first and fifth toes were not included because of several competitive nail involvements mimicking psoriasis (fungal infection, injury, etc.).

In case of doubt about nail involvement, photos could be sent to a dermatology consultant for advice.

**PATIENT-REPORTED OUTCOMES SPECIFICATIONS:**

The patients entered the PROs before study inclusion as part of routine care in the outpatient clinic. A consultation could be a physician or nurse consultation, infusion with bDMARD, or pick-up of medicine (DMARD). PRO entries should be a maximum of seven days old at inclusion.

**TRANSLATION OF THE PATIENT-REPORTED OUTCOME MEASURES:**

We engaged two independent translators, both native speakers of Danish and English, to translate the questions from Danish to English. One had a medical background, while the other did not. Using the two translations, we created a consensus version, which is presented in Table 1.

**STUDY PHYSICIAN COMPETENCIES**:

The physician had six years of clinical experience, including one year in rheumatology. Additionally, the study physician had received training from an experienced dermatologist in assessing skin- and nail psoriasis and spent time mastering the Leeds Dactylitis Index[1].

Supplementary Figure S1: Flow chart of participants

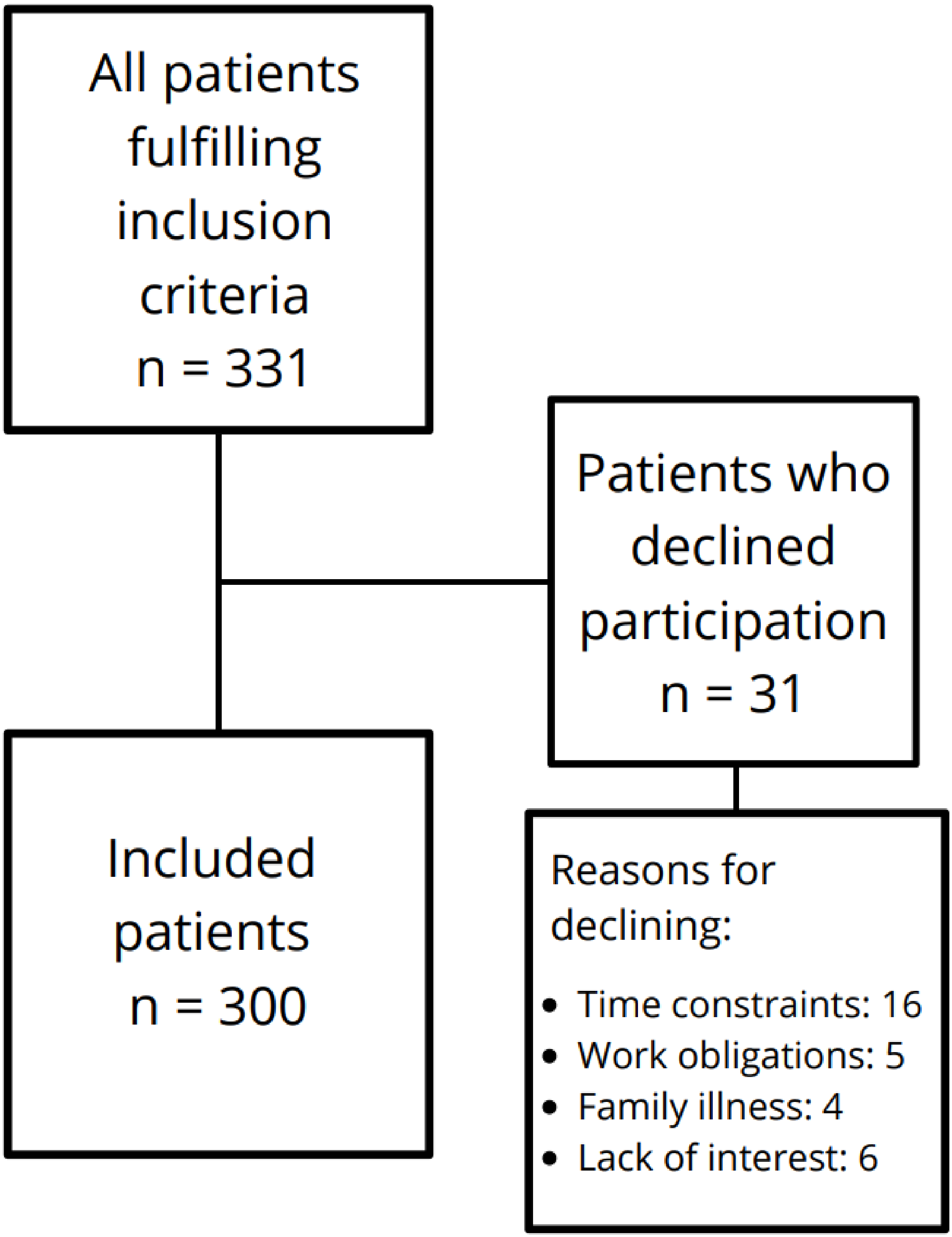

### Supplementary Table S1

Crosstabulation of patient-reported outcomes and physician assessments for the PROMs covering dactylitis, skin psoriasis, nail psoriasis, and uveitis before (Step 1) and after (Step 2-3) the added dactylitis photo and before (Step 2) and after blinding of the physician (Step 3).

#### Dactylitis, n=300

**Dactylitis before photo n = 134 patients (Step 1)** — Study physician assessment

| | | Yes | No | Do not know |
|---|---|---|---|---|
| PRO | Yes | 9 | 24 | 1 |
| | No | 3 | 89 | 0 |
| | Do not know | 1 | 7 | 0 |

**Dactylitis after photo n = 166 patients (Step 2-3)** — Study physician assessment

| | | Yes | No | Do not know |
|---|---|---|---|---|
| PRO | Yes | 23 | 20 | 0 |
| | No | 3 | 101 | 0 |
| | Do not know | 2 | 17 | 0 |

#### Dactylitis, after photo (Step 2-3), n= 166

**Dactylitis before blinding n = 66 patients (Step 2)** — Study physician assessment

| | | Yes | No | Do not know |
|---|---|---|---|---|
| PRO | Yes | 7 | 6 | 0 |
| | No | 1 | 46 | 0 |
| | Do not know | 1 | 5 | 0 |

**Dactylitis after blinding n = 100 patients (Step 3)** — Study physician assessment

| | | Yes | No | Do not know |
|---|---|---|---|---|
| PRO | Yes | 16 | 13 | 0 |
| | No | 2 | 56 | 0 |
| | Do not know | 1 | 12 | 0 |

#### Psoriasis, skin and nails, n=300

**Skin psoriasis before blinding = 200 (Step 1-2)** — Study physician assessment

| | | Yes | No | Do not know |
|---|---|---|---|---|
| PRO | Yes | 109 | 4 | 2 |
| | No | 1 | 80 | 0 |
| | Do not know | 0 | 4 | 0 |

**Skin psoriasis after blinding = 100 (Step 3)** — Study physician assessment

| | | Yes | No | Do not know |
|---|---|---|---|---|
| PRO | Yes | 54 | 2 | 3 |
| | No | 0 | 32 | 0 |
| | Do not know | 0 | 6 | 3 |

**Nails psoriasis before blinding = 200** — Study physician assessment

| | | Yes | No | Do not know |
|---|---|---|---|---|
| PRO | Yes | 78 | 4 | 0 |
| | No | 17 | 83 | 1 |
| | Do not know | 6 | 11 | 0 |

**Nail psoriasis after blinding = 100** — Study physician assessment

| | | Yes | No | Do not know |
|---|---|---|---|---|
| PRO | Yes | 45 | 2 | 0 |
| | No | 14 | 34 | 1 |
| | Do not know | 3 | 0 | 1 |

#### Uveitis n= 300

**Uveitis (Step 1-3)** — Study physician assessment

| | | Yes | No | Do not know |
|---|---|---|---|---|
| PRO | Yes | 3 | 4 | 0 |
| | No | 0 | 292 | 0 |
| | Do not know | 0 | 1 | 0 |

The grey-scaled areas are the cases where the patient and the physician agree.

**Abbreviation**: Patient-reported outcome measures (PROM), Patient-reported outcome (PRO)
